# DNA Methylation Signatures of Alcohol Use Disorder – A large-scale Meta-Analysis in the Psychiatric Genomics Consortium

**DOI:** 10.1101/2025.08.29.25334705

**Authors:** Lea Zillich, Sofia D’Augello, Diana Avetyan, Graciela E. Delgado, Ian R. Gizer, Jeesun Jung, Seyma Katrinli, Marcus E. Kleber, Natalie Merrill, Angela P. Moissl, Diana L. Núñez-Rios, Jacqueline M. Otto, Eric Zillich, Eva Friedel, Dana B. Hancock, Eric O. Johnson, José Jaime Martínez-Magaña, Sheila T. Nagamatsu, David W. Sosnowski, Julie D. White, Karolina A. Aberg, William E. Copeland, Nancy Diazgranados, Negar Fani, Joel Gelernter, David Goldman, Jerome C. Foo, Henry R. Kranzler, Daniel F. Levey, Winfried März, Brenda W. J. H. Penninx, Renato Polimanti, Abigail Powers, Alicia K. Smith, Rainer Spanagel, Edwin J. C. G. van den Oord, Kirk C. Wilhelmsen, Stephanie H. Witt, Katharina Domschke, Miriam A. Schiele, Brion S. Maher, Janitza L. Montalvo Ortiz, Shaunna L. Clark, Falk W. Lohoff

**Author notes:** Corresponding Author: Dr. Lea Zillich, University Medical Center Freiburg, Department of Psychiatry and Psychotherapy, Hauptstr. 5, 79104 Freiburg, Germany. authors contributed equally (Shaunna L. Clark, Falk W. Lohoff).

## Abstract

Despite extensive research on DNA methylation (DNAm) signatures associated with alcohol use disorder (AUD), findings are often inconsistent and not replicated. We conducted a large-scale meta-analysis of epigenome-wide association studies (EWAS) to identify reliable, reproducible epigenetic markers of AUD.

Seven cohorts, comprising 3,775 individuals (1,325 with AUD), contributed to this meta-analysis within the framework of the Psychiatric Genomics Consortium Substance Use Disorders Epigenetics Working Group. Downstream analyses included the identification of differentially methylated regions, overrepresentation analyses, and the construction of a methylation risk score (MRS).

We identified 118 significant CpG sites associated with AUD, with the strongest association found at cg24889777 (*p*=5.12×10^-17^) in the long non-coding RNA LOC100505942. CpG sites were enriched for pathways related to GTPase signaling and transmembrane transporter activity, as well as EWAS signals of alcohol consumption. The MRS explained 10.44% of variance in heavy drinking in an independent cohort (N=2,534, AUC=0.657).

This large-scale meta-analysis offers key insights into the epigenetic mechanisms of AUD and lays the groundwork for future research on methylation risk scores for the diagnosis, prognosis, and treatment in AUD.

## Introduction

Alcohol Use Disorder (AUD) is characterized by unhealthy alcohol use and includes multiple clinical symptoms, such as the loss of control over alcohol intake, the development of tolerance, and withdrawal symptoms when alcohol is not consumed^1^. AUD is a heterogeneous phenotype, with affected individuals exhibiting a wide variety of alcohol use behaviors that are associated with different clinical, social, and economic correlates^2,3^. At the same time, AUD is a common disorder that affects about 10% of the population in a given year and is associated with significant morbidity and mortality^4^. Despite the negative health effects, treatment options for AUD are limited, and there is no clear understanding of the underlying neurobiology^5^.

Recently, there has been an increasing interest in how epigenetic factors contribute to the risk and pathology of AUD^6,7^. Epigenetic modifications are reversible changes that can influence gene function without altering the DNA sequence itself, thus serving as vital and potentially mechanistically informative genetic and environmental indicators of gene/environment interaction^8^. This is particularly relevant for AUD, for which the heritability is estimated to be about 50% based on twin studies^9^. While genome-wide association studies (GWAS) can reflect the risk of developing an AUD, epigenome-wide association studies (EWAS) additionally capture the effects of alcohol exposure, next to other environmental and lifestyle factors, and could contribute to disentangling the heterogeneity in AUD. Epigenetic modifications involve various mechanisms, with DNA methylation being the most studied in humans. DNA methylation primarily involves the addition of a methyl group to the 5’ position of cytosines and is typically observed in cytosine-guanine dinucleotides (CpG sites)^8^. DNA methylation is highly tissue- and cell type-specific. While it is primarily metabolized in the liver, alcohol has systemic effects on the entire organism^10^. Because of the tissue-specificity of DNA methylation in combination with the widespread effects of alcohol, investigating DNA methylation in peripheral blood offers the opportunity to capture these systemic effects. At the same time, there is high translational value in investigating peripheral DNA methylation, which allows for the construction of methylation scores that have potential as biomarkers, as demonstrated for aging^11^, alcohol consumption^12^, and smoking^13^. A similar marker, based on robust DNA methylation associations, could plausibly be constructed for AUD^14^.

There have been several epigenome-wide association studies (EWAS) investigating the association between DNA methylation and AUD phenotypes in peripheral blood samples^15-17^. However, small sample sizes ranging between 198 and 1,132, and the resulting limited statistical power, have produced inconsistent results, but also provided the basis for a larger investigation across cohorts. So far, replicated CpG sites have been observed in *GAS5*, which was identified as a target for epigenetic modification in an EWAS comparing 379 individuals with and 246 without AUD in the National Institute on Alcohol Abuse and Alcoholism (NIAAA) cohort^15^. This site was also replicated in an independent sample, the Grady Trauma Project (GTP). The largest EWAS to date included 323 individuals with AUD and 809 unaffected controls in the Netherlands Study of Depression and Anxiety (NESDA)^16^. Here, *DLGAP1* was identified and confirmed in 412 participants of the Great Smoky Mountains Study (GSMS)^16^. In addition to the need to increase sample sizes, it is important to investigate DNA methylation associated with AUD across heterogeneous cohorts to identify robust signals representative of the heterogeneity of the phenotype regarding severity, comorbidities, and somatic factors, among others. The currently available studies range from cohorts ascertained for AUD to population-based approaches and cohorts originally ascertained for comorbid disorders, such as major depressive disorder (MDD) and posttraumatic stress disorder (PTSD). Large-scale meta-analyses or large samples are necessary to facilitate valid and replicable results that are independent of specific cohort profiles. The advantages of meta-analysis are increasing the sample size, enhancing statistical power, and including cohorts with different characteristics; a disadvantage is heterogeneity that may be hard to identify or account for statistically.

The present study aimed to identify DNA methylation signals robustly associated with AUD, construct a methylation risk score (MRS), and perform out-of-sample predictions. To accomplish this, we first conducted the largest meta-analysis of epigenome-wide association studies (EWAS) on AUD to date, investigating seven cohorts with a total sample size of 3,775 individuals (AUD = 1,325, controls = 2,450). Next, we carried out extensive downstream analyses, including overrepresentation analysis of Gene Ontology terms and genes identified in genome-wide association studies (GWAS) to explore potential functional mechanisms behind the findings. We also tested the overlap with alcohol consumption and the blood-brain concordance and performed sensitivity analyses to control for smoking effects. Finally, we constructed a methylation risk score and validated it in an independent cohort (N=2,534).

## Results

### Meta-analysis of alcohol use disorder

To identify robust associations of DNA methylation and AUD, a meta-analysis of standardized, cohort-level EWASs was performed, resulting in a sample size of 3,775 individuals of primarily European Ancestry (Table 1). Participating cohorts included 194 male patients with AUD, recruited at the Central Institute of Mental Health (CIMH) in Mannheim^17^, 231 participants from the Yale-Penn cohort^18,19^, 442 participants from the University of California at San Francisco Family Alcoholism Study (UCSF)^20^, 677 individuals who participated in the Grady Trauma Project (GTP)^21^, 615 patients who were treated at the National Institute on Alcohol Abuse and Alcoholism (NIAAA)^15^, 484 participants of the Great Smoky Mountains Study (GSMS)^22,23^, and 1,132 participants of the Netherlands Study on Depression and Anxiety (NESDA)^24,25^. Detailed information on the cohorts can be found in Supplementary Text S1. The main model tested the association of DNA methylation with AUD, including covariates for sex, age, self-reported smoking, cell type composition, and three genotype principal components to control for population stratification. All array-based EWASs also included four principal components of the internal control probes to control for batch effects. The Quantile-Quantile (QQ) plot and lambda value (λ = 1.28) indicated a modest degree of inflation (Extended Data 1). Bacon correction was applied to control inflation, resulting in a final lambda of 1.06.

| Cohort | N samples | N case | N control | DNA<br>Methylation<br>Measurement | self-reported<br>Ancestry | CpGs | Lambda |
| --- | --- | --- | --- | --- | --- | --- | --- |
| **CIMH,**<br>**Germany** | 194 | 99 | 95 | EPIC-Array v1 | European | 700163 | 1.264 |
| **Yale-Penn,**<br>**USA** | 231 | 75 | 156 | EPIC-Array v1 | European | 702116 | 0.859 |
| **UCSF**<br>**Family**<br>**Alcoholism**<br>**Study, USA** | 442 | 243 | 199 | 450K-Array | European | 320910 | 0.923 |
| **GTP, USA** | 677 | 52 | 625 | EPIC-Array v1 | African<br>(95%) | 672278 | 1.007 |
| **NIAAA,**<br>**USA** | 615 | 372 | 243 | EPIC-Array v1 | European/<br>African<br>American | 703800 | 1.127 |
| **GSMS, USA** | 484 | 161 | 323 | MBD-seq | European/<br>African<br>American/<br>Native<br>American | 586902 | 0.988 |
| **NESDA,**<br>**Netherlands** | 1132 | 323 | 809 | MBD-seq | European | 5632173 | 1.03 |
| **TOTAL** | **3775** | **1325** | **2450** |  |  |  |  |

CIMH = Central Institute of Mental Health, UCSF = University of California San Francisco Family Alcoholism Study, GTP = Grady Trauma Project, NIAAA = National Institute on Alcohol Abuse and Alcoholism, GSMS = Great Smoky Mountains Study, NESDA = Netherlands Study of Depression and Anxiety, MBD-Seq = Methyl-Binding-Domain Sequencing, EA = European Ancestry, AA = African Ancestry, NAA = Native American Ancestry

In the fixed-effects meta-analysis, 118 CpG sites were significantly associated with AUD after Bonferroni correction; see also Fig. 1 and Supplementary Table S1. Genome-wide CpG methylations are non-independent. However, overall patterns of covariance are not well understood and are likely to be affected by tissue, developmental timing, and exposure. Thus, for the purposes of this analysis, a conservative threshold for significance, assuming independence of the CpGs assayed, was used. The strongest association was observed for cg24889777 (B_bacon_=0.021, *p*_bacon_=5.12×10^-17^) in the long non-coding RNA LOC100505942. The top CpG sites showed consistent effects across cohorts, which are depicted in Fig. 1B-G for the six most significant CpG sites exhibiting converging effects in at least five of the seven cohorts. Each of these was also directionally consistent.

**Figure 1.**
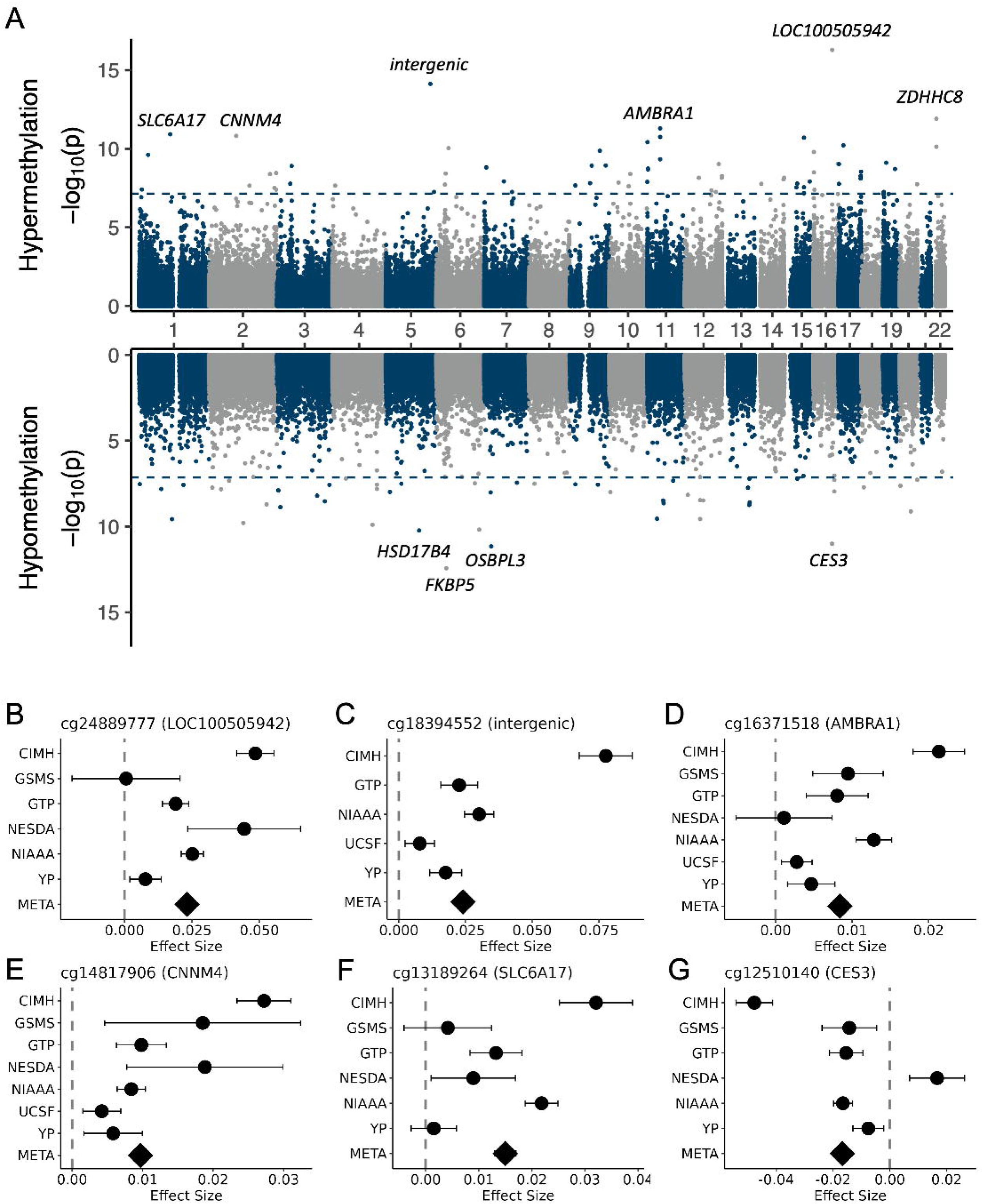
A) Miami Plot depicting differentially methylated positions (DMPs) split into hypermethylated (upper panel) and hypomethylated (lower panel) CpG sites; genes of the ten most significant DMPs are annotated; blue dashed line indicates epigenome-wide significance. B)-G) regression coefficients for AUD for the six most significant CpG sites in the meta-analysis with converging effect estimates in at least five cohorts; error bars represent standard errors of estimate, diamond shape represents effect in meta-analysis; META = primary meta-analysis, YP = Yale-Penn, UCSF = University of California at San Francisco, NIAAA = National Institute on Alcohol Abuse and Alcoholism, NESDA = Netherlands Study of Depression and Anxiety, GTP = Grady Trauma Project, GSMS = Great Smoky Mountain Study, CIMH = Central Institute of Mental Health; red dashed line indicates effect size in the meta-analysis.

To evaluate technical effects due to DNA methylation assessment, we conducted an additional analysis in which cohorts evaluating DNA methylation via microarray or MBD-seq were meta-analyzed separately. This was followed by a second meta-analysis. The correlation between the effect sizes of the microarray analysis and the primary analysis was *r* = 0.99 (*p* < 2.2×10^-16^), while the correlation between the MBD-seq results and the primary analysis was *r* = 1 (*p* < 2.2×10^-16^); see also Extended Data 2A-B.

In addition, 57 significantly differentially methylated regions (DMRs) were identified after false discovery rate (FDR) correction (Supplementary Table S2 and Extended Data 2C). The top significant DMR was in *HNRNPA1* (p_FDR_ = 1.62×10^-17^), while the DMR containing the most significant CpG site was annotated in the *AMBRA1* gene (p_FDR_ = 1.43×10^-15^).

#### Gene Ontology Overrepresentation Analysis

The 118 epigenome-wide significant CpG sites were enriched for biological processes related to amino acid transport, such as “L-leucine transport” (GO:0015820, z = 14.26, *p*_FDR_ = 0.016) and “branched-chain amino acid transport” (GO:0015803, z = 11.98, *p*_FDR_ = 0.023), molecular functions related to GTPase binding and transmembrane transporter activity, as well as cellular components related to vesicle transport, such as “transport vesicle” (GO:0030658, z = 5.79, *p*_FDR_ = 0.0063) and “phagocytic vesicle” (GO:0045335, z = 6.94, *p*_FDR_ = 0.0063), as well as membrane-related GO terms.

As with CpG sites, DMRs were enriched for membrane-related cellular components and molecular functions related to GTPase binding and transmembrane transporter activity. Enrichment was also observed for biological processes related to response to nutrient levels, such as “cellular response to starvation” (z = 8.43, *p*_FDR_ = 0.005). Results are visualized in Extended Data 3, and all nominally significant GO terms for CpG sites are summarized in Supplementary Tables S3 and S4 for DMRs.

#### GWAS Enrichment Analysis

To explore whether genetic variance contributed to the findings, we formed three gene sets consisting of: CpG sites identified in the primary meta-analysis, CpG sites identified in the primary meta-analysis with known methylation quantitative trait loci (meQTLs)^26^, and DMRs. While none of these gene sets were significantly enriched for GWAS signals of problematic alcohol use^27^, alcohol dependence^28^, drinks per week^29^, cigarettes per day^30^, MDD^31^, and PTSD^32^ (all *p* > 0.056; Supplementary Table S5), CpG sites in DMRs showed the strongest associations with GWAS signals for alcohol dependence^28^ (*p* = 0.056) and drinks per week^29^ (*p* =0.073).

#### Overlap with Alcohol Consumption EWAS

Because alcohol consumption is a necessary prerequisite to AUD, we investigated whether the DNA methylation signatures of AUD were distinct or shared with those of alcohol consumption traits. Therefore, we performed an overlap analysis with the results of a recent EWAS of alcohol consumption in the Generation Scotland cohort^33^. Of the 118 epigenome-wide significant CpG sites identified in the primary analysis, 65 (55.1%) were also significantly associated with alcohol consumption, indicating a substantial overlap between DNA methylation signatures of alcohol consumption and AUD (*p* < 2.2×10^-16^), see also Fig. 2A. The direction of effect was the same for all overlapping CpG sites (Fig. 2B). Two of the top ten CpG sites were exclusively associated with AUD, namely the top hit, cg24889777, and cg03546163 in *FKBP5*.

**Figure 2.**
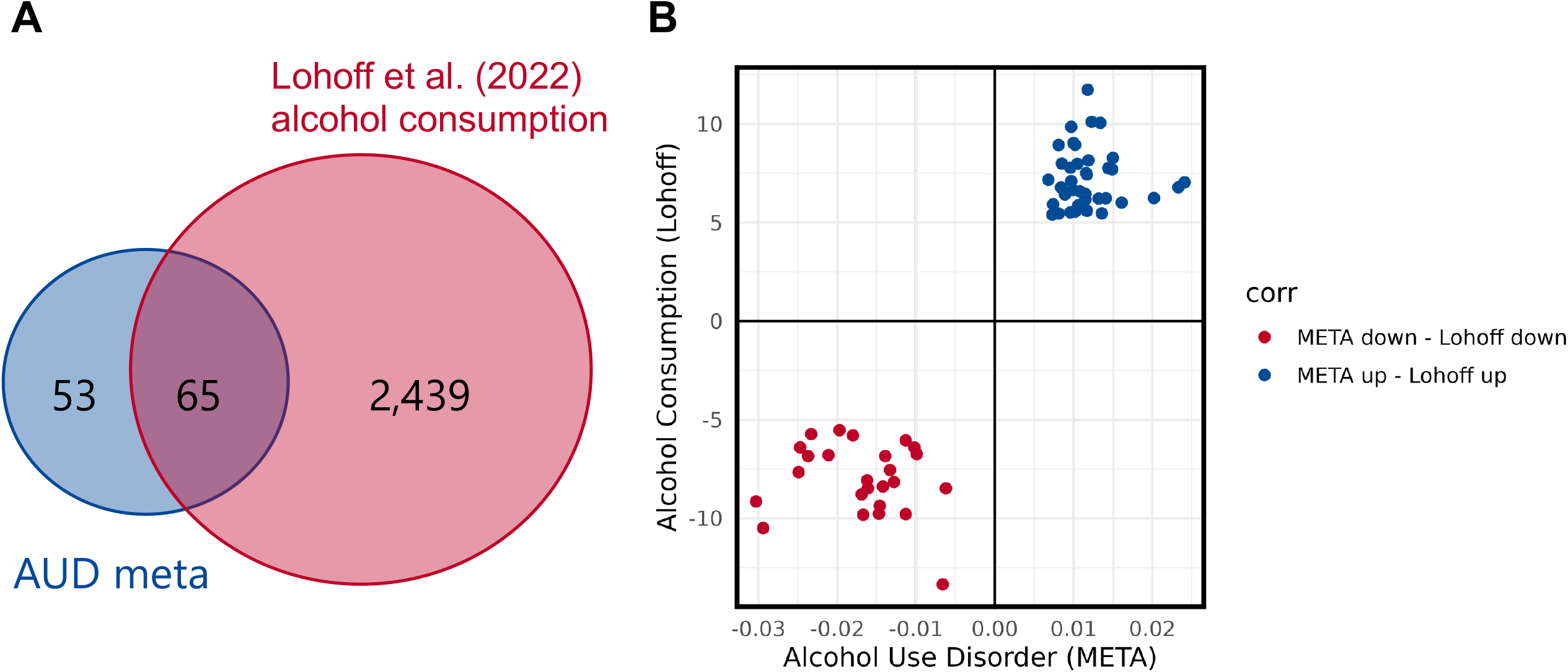
A) Venn Diagram depicting the overlap between the primary meta-analysis and CpG sites significantly associated with alcohol consumption in Lohoff et al. (2022). B) Scatterplot depicting the directions of effects in Lohoff et al. (2022) and the primary meta-analysis.

#### Blood-brain concordance

Due to the tissue-specificity of DNA methylation, we investigated the correlation between the DNA methylation levels of the identified CpG sites in blood and brain tissue. First, we investigated whether a correlation between blood and brain DNA methylation levels of the top 100 CpG sites associated with AUD in the primary meta-analysis could be observed using BECon^34^. This analysis revealed information on blood-brain concordance for 33 CpG sites. The full results are presented in Extended Data 4. Five CpG sites exhibited strong correlations (0.49 ≤ *r* ≤ 0.60) in either Brodmann Area 10, i.e., cg06846976 in *TSC2* and cg04406114 in *POU2F2*, in Brodmann Area 20, i.e., cg14380013 in *UNC119B* and cg03546163 in *FKBP5*, or Brodmann Area 7, i.e., cg18059012 in *PATL2*.

In addition, we investigated whether the 118 CpG sites identified in the primary meta-analysis overlapped with results from two recent EWASs of AUD in postmortem human brain tissue from Zillich et al. (2022)^35^ and White et al. (2024)^36^. In the two studies, five brain regions that play a central role in the neurocircuitry of addiction were investigated: dorsolateral prefrontal cortex/Brodmann Area 9^35,36^, ventral striatum/nucleus accumbens^35,36^, caudate nucleus^35^, putamen^35^, and anterior cingulate cortex^35^. There was no overlap between the CpG sites identified in the primary meta-analysis and the postmortem brain EWASs.

#### Smoking Sensitivity Analysis

Results from the sensitivity analysis in nonsmokers largely overlapped with those of the primary analysis. All 118 significant CpG sites in the primary meta-analysis were also at least nominally significant in the nonsmoking analysis. The effect sizes of the nominally significant CpG sites in the primary analysis were strongly associated with those in the nonsmoking analysis (*r* = 0.88, *p* < 2.2×10^-16^); see also Extended Data 5.

### Methylation Risk Score

For a potential clinical translation, we constructed an MRS based on the 118 epigenome-wide significant CpG sites identified in the primary meta-analysis and validated the score in the independent LURIC cohort. The LURIC (Ludwigshafen Risk and Cardiovascular Health Study) cohort was recruited between 1997 and 2000 and included 3,316 patients who were hospitalized for a coronary angiography^37^. For 2,534 of these patients, DNA methylation data and detailed information on alcohol consumption were available. Patients reported their mean daily intake of alcoholic beverages, which was then converted to grams of alcohol consumed per day as described in Moissl et al. (2021)^38^. While AUD diagnosis was not assessed in LURIC, the cohort includes a large number of participants (37.05%) exhibiting heavy drinking according to NIAAA criteria (Fig. 3A)^39^.

**Figure 3.**
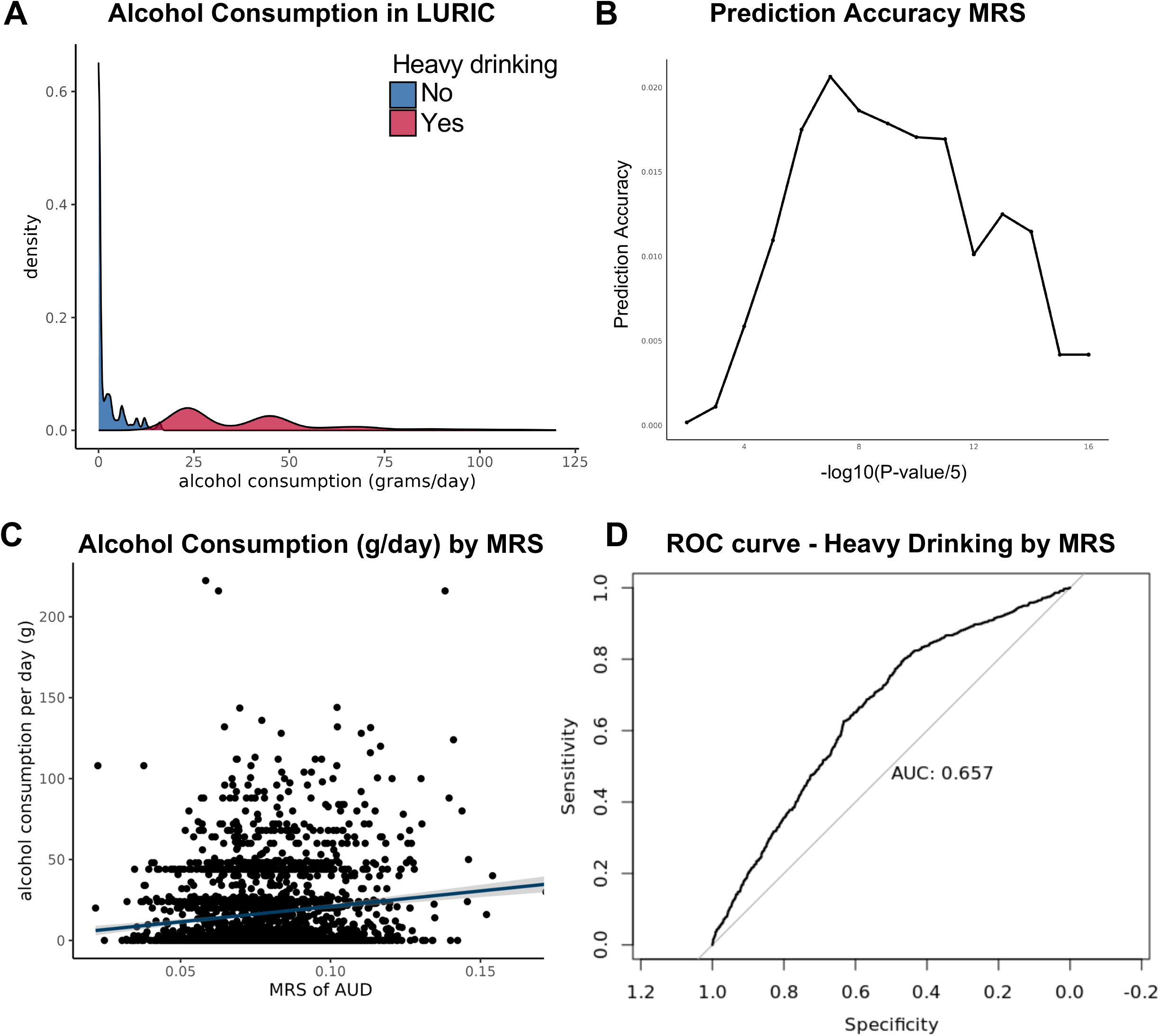
A) alcohol consumption in LURIC by heavy drinking status, red=heavy drinking, blue= no heavy drinking, B) Prediction accuracy (R^2^) of the methylation risk score (MRS) of AUD for p-value thresholds, C) alcohol consumption per day in grams, predicted by the MRS of AUD, D) ROC curve for heavy drinking.

MRS were constructed using the pruning and thresholding method proposed by Chen et al (2023)^40^. Here, methylation values were first combined into co-methylated regions and, based on different p-value thresholds, lead CpG sites per region were identified. The AUD MRS was then constructed based on the weights of the lead CpG sites. The p-value thresholding resulted in a p-value threshold of 5×10^-7^, as presented in Fig. 3B. The resulting AUD MRS comprised 204 CpG sites after pruning and explained 2.06% of the variance in alcohol consumption (grams/day) in the LURIC cohort. When investigating the MRS in linear regression models, controlling for sex, age, and cell type composition, the AUD MRS remained predictive of alcohol consumption (B=5.24, SE=0.59, p<2×10^-16^). In addition to the AUD MRS, sex, age, and CD8^+^- and CD4^+^ T-cells were predictive of daily alcohol consumption (Supplementary Table S6). The overall model explained 10.44% of the variance.

Using logistic regression, we found that the AUD MRS was predictive of heavy drinking (B=6.77, SE=1.33, p<2×10^-16^), serving as a potential proxy for AUD. Sex and the CD8^+^ T-cell proportion were also significant predictors in the model, although with smaller effects (see also Supplementary Table S7 and Fig. 3C). Nagelkerke’s R^2^ showed that the model accounted for 10.48% of the variance in heavy drinking, with an area under the receiver operating characteristic curve of 0.657 (Fig. 3D).

## Discussion

The present meta-analysis identified several differentially methylated CpG sites and regions robustly associated with AUD. Our results highlight CpG sites showing novel associations with AUD, as well as genes previously associated with alcohol consumption and related phenotypes, and reproduced findings from previous studies^15-17,33^. We also constructed an MRS of AUD that predicted alcohol consumption and heavy drinking in an independent cohort.

The most significant DMP was cg24889777 in the locus *LOC100505942*, which transcribes an uncharacterized non-coding RNA. This CpG site was also associated in previous EWAS of smoking and COPD, incident liver cirrhosis, chronic pain, prevalent ischemic heart disease, and incident type 2 diabetes^41,42^; phenotypes that show a strong genetic and phenotypic association and clinical overlap with AUD^43-46^. Differential methylation in cg24889777 is a novel finding in AUD and has not previously been associated with alcohol consumption. The association with traits and disorders comorbid with AUD suggests that methylation differences in cg24889777 could reflect shared underlying mechanisms between these traits that could affect non-coding RNA regulation.

Several CpG sites previously linked to AUD in individual studies were also identified in the primary meta-analysis, underlining the robustness of associations. For example, *SLC1A2*, which encodes the glutamate transporter 1, was robustly associated in the meta-analysis and in prior analyses^15,17^. Other genes include *HNRNPA1* and *AMBRA1*, both of which emerged in the positional and regional analyses. *AMBRA1* encodes the autophagy and beclin 1 regulator 1, responsible for GTPase binding and ubiquitin protein ligase binding activity. It is expressed in various tissues, including the brain. As with the top DMP, the CpG site cg14222701, located in the *AMBRA1* gene, was differentially methylated in previous EWASs of type 2 diabetes, chronic pain, smoking, COPD, lung cancer, and ischemic heart disease^41,42^. In two studies, differential methylation in *AMBRA1* was associated with AUD^17^ or alcohol consumption^12^. Together with the DCX (DDB1-CUL4-X-box) E3 ubiquitin-protein ligase, AMBRA1 assembles into a complex that plays a role in the cell cycle and autophagy^47^. The substrates targeted by the DCX(AMBRA1) complex were previously associated with alcohol consumption. One example is the *CND* gene coding for Cyclin D, a polyubiquitination target^47^. In one study, different CCNDs were differentially expressed in the hippocampus of people with AUD, revealing pathways responsible for cell cycle regulation or cancer signaling to be associated with AUD^48^. The autophagosomal marker beclin 1, another target of the DCX(AMBRA1) complex, was increased in association with alcohol exposure and is thought to play a role in ethanol-induced autophagy^49^.

We observed a substantial overlap between DNA methylation signatures of AUD and those of alcohol consumption^33^. While alcohol consumption is a prerequisite of AUD, the considerable overlap could indicate that some of the observed DNA methylation signatures reflect the prolonged alcohol consumption associated with AUD. In line with this, alcohol consumption and AUD also show both shared and distinct genetic architectures, although GWAS signals are thought to primarily reflect vulnerability, while EWAS also capture the consequences of exposure. At the same time, our results did not indicate a strong enrichment of the results of the primary meta-analysis for GWAS signals of problematic alcohol use, alcohol dependence, or drinks per week, even though trends were observed for DMR enrichment for the latter two traits. To disentangle the genetic and epigenetic signatures of AUD, both polygenic and methylation risk scores can prove useful. In the present study, the MRS derived from the primary analysis predicted heavy drinking in an independent cohort. The performance of the MRS was similar or slightly higher than those recently reported for other psychiatric phenotypes, such as major depressive disorder (AUC-ROC = 0.53)^50^ or for a childhood trauma MRS predicting psychopathology 17 years later in early adulthood (0.54 ≤ AUC-ROC ≤ 0.63)^23^. Previous studies that developed MRS of AUD did not test them in independent samples, or the datasets used to construct the MRS were from small samples^14^. Here, we construct an MRS predictive of alcohol consumption and heavy drinking as a binary phenotype. At the same time, due to lack of available data, the performance of the MRS was not tested in an independent sample comparing individuals with and without an AUD diagnosis. Nevertheless, the current analysis provides an important basis for translational research and testing the clinical applications of the AUD MRS in independent cohorts.

While some of the identified CpG sites showed blood-brain correlations, little to no overlap was observed with findings from recent EWAS of AUD in postmortem human brain tissue^35,36^. This could be due to the small sample size in the human postmortem EWAS, which ranged from 46 to 119^35,36^. This highlights the importance of combining datasets to enhance statistical power and facilitate the replication of individual study results. Currently, many of the findings from EWASs of AUD in postmortem human brain tissue have not been replicated^16,35,36,51^, necessitating similar meta-analytical approaches to identify robust findings for which, in turn, the overlap with peripheral blood markers can be determined.

While we present the most extensive DNA methylation meta-analysis to date, the sample size and resulting statistical power remains a potential limitation. In addition, we cannot exclude confounding factors not accounted for in the present analysis that could have driven the associations observed between AUD and DNA methylation. We show in sensitivity analyses that one major confounding factor, smoking, did not seem to influence our results, as demonstrated by a strong overlap between the nonsmoking analysis and the primary meta-analysis. Due to the correlational nature of the analysis, it is unclear whether DNA methylation at the identified CpG sites is altered because of heavy drinking associated with AUD or whether the DNA methylation itself contributes to the risk of developing AUD. Another limitation is the analysis of bulk tissue that can mask cell type-specific effects of DNA methylation. While we controlled for the major cell type proportions in whole blood, future studies on the single-cell level or using sorted cell populations should be performed to directly investigate cell type-specific DNA methylation signatures of AUD. While we can make assumptions on how differential DNA methylation might influence biological processes through pathway analyses, future studies should also investigate how DNA methylation signatures of AUD affect other -omics signatures, such as transcriptomics.

Taken together, we identified differentially methylated CpG sites and regions robustly associated with AUD in more than 3,700 participants across seven cohorts representing heterogeneous AUD phenotypes. Our results identified new targets and reproduced findings from previous EWAS. The resulting MRS was predictive of heavy drinking in an independent cohort, underscoring the potential for use of the MRS in research and eventually in the clinic.

## Methods

### Samples

DNA methylation data from peripheral blood samples of a total of 3,775 individuals, 1,325 individuals with and 2,450 without AUD, from seven cohorts, all members of the Substance Use Disorder Epigenetics Working Group of the Psychiatric Genomics Consortium (PGC), were included in the meta-analysis. Three cohorts explicitly focused on individuals with AUD: the CIMH cohort (N=194) followed male patients who underwent withdrawal treatment for AUD and matched controls^17^; in the UCSF Family Alcoholism Study (N=442), families with an increased risk for AUD were investigated^20^, and the NIAAA cohort investigated patients seeking treatment for AUD at the NIAAA (N=615)^15^. The Yale-Penn cohort (N=231)^18,19^ included subjects recruited because they had substance dependence traits or were controls and the Grady Trauma Project (N=650)^21^ focused on veterans with posttraumatic stress disorder, which is highly comorbid with substance use disorders, such as AUD. Similarly, the NESDA study (N=1,132) recruited participants with depression and anxiety^25^, also highly comorbid with AUD. GSMS (N=540)^22^ is a longitudinal study from early childhood to adulthood enriched for participants with a higher risk of developing an SUD, though only adult time points are considered here. Additional cohort characteristics are described in the Supplementary Material. In all cohorts, AUD was diagnosed according to the Diagnostic and Statistical Manual of Mental Disorders (DSM) version IV (alcohol dependence) or version 5 (at least moderate alcohol use disorder).

### DNA Methylation

DNA methylation levels were determined in five cohorts using the Illumina Infinium Human Methylation EPIC BeadChip v1.0 (about 850,000 probes) or the 450K BeadChip (about 450,000 probes). In NESDA and GSMS, Methyl-binding domain (MBD) sequencing was performed.

### Quality Control of raw data

A standardized quality control pipeline was distributed among all cohorts with microarray data, including the replication cohort LURIC, a customized version of the CPACOR pipeline published by Lehne, et al.^52^. Raw IDAT files were read and processed using the *minfi* package. Illumina background subtraction was performed to remove outliers by removing the average negative control signal intensity. A principal component analysis (PCA) of the positive control probe intensities was performed to remove technical biases in the EWAS. Probes were excluded if the detection p-value was larger than 10^-16^, resulting in a marker call rate lower than 95%, if the beadcount was lower than three in more than 5% of samples, and if probes were located on the sex chromosomes. Samples were removed if the sample call rate was below 95% or if the reported sex did not match biological sex. Cell type composition was estimated using the EpiDISH package^53^, resulting in estimates for CD8T cells, CD4T cells, natural killer cells (NK), B cells (B), monocytes (mono), and neutrophils.

The quality control procedure for MBD-seq is described in detail in Clark, et al.^16^. Briefly, sequence reads were aligned to the human reference genome (hg19/GRCh37) using Bowtie2^54^. Quality control of samples, reads, and sites was conducted using RaMWAS^55^ as described elsewhere^25,56^ and excluded samples with failed enrichment or if reported sex did not match biological sex, resetting read counts if there were excessive duplicate reads (>3 reads starting at the same location were reset to 1), excluding reads in difficult to align loci, and, excluding sporadically methylated sites or those located on sex chromosomes. Using a non-parametric estimate of fragment size distribution, methylation scores were calculated by estimating the number of fragments covering each CpG. These scores represent a quantitative measure of methylation for each sample at a specific CpG. Cell type composition was estimated using Houseman’s method^57^ and a reference set of methylomes based on MBD-seq data described elsewhere^16^, resulting in estimates for CD15 granulocytes, CD3T cells, B cells, and monocytes.

### Cohort-level EWAS

A standardized analysis plan was distributed among participating cohorts. All cohorts with microarray DNA methylation data were additionally provided with a standardized analysis pipeline, while the MBD-seq cohorts ran similar models based on the analysis plan. Before running the EWAS, all variables were tested for multicollinearity by calculating Pearson correlations and flagging variable pairs with a correlation coefficient greater than 0.70. For the cohort-level EWAS, DNA methylation beta values were predicted by AUD status in robust linear models using the R package MASS. All cohorts controlled for age, smoking, cell type proportions (CD8T, CD4T, NK, B, mono), three genotype principal components, and four principal components of the internal control probes. Cohort-specific covariates were sex and covariates based on cohort ascertainment, such as MDD, PTSD diagnosis. Detailed descriptions of cohort specificities are provided in Supplementary Text S1.

### Quality Control and Filtering of EWAS summary statistics

We performed additional filtering for cross-reactive probes and SNPs in base extensions with a minor allele frequency > 1% in European and African American Ancestry^55^. We used the EWAS_QC function of the R package QCEWAS^56^ to calculate lambda values for the cohort results, inspect the distribution of estimates and standard errors, and explore individual-level results. For the primary meta-analysis, MBD-seq results were restricted to CpG sites on the EPIC BeadChip, and the number of sites included is presented in Table 1.

### Meta-Analysis

The quality-controlled and filtered CpG sites were utilized for an invariance-weighted fixed-effects meta-analysis using METAL (version 2011)^58^. The meta-analysis focused on CpG sites in at least N=1,929 samples or at least 50% of the samples, resulting in 25,160 skipped and 708,385 analyzed CpG sites. In addition to the primary meta-analysis, we performed secondary analyses based on methylation quantification, MBD-seq and microarray. These results were subsequently meta-analyzed, and correlation analysis investigating the convergence between the effect sizes of the primary meta-analysis and the secondary meta-analysis was performed. All results were corrected for multiple testing using the Bonferroni correction. Results were annotated using the manufacturer’s manifest and visualized using the miamiplot R package^59^.

### Downstream Analyses

#### Differentially Methylated Regions

Differentially methylated regions (DMRs) were identified using the comb-p algorithm^60^, as implemented in ENmix (version 1.42.2)^61^. Comb-p can be used solely on summary statistics and is therefore well-suited for meta-analysis results. All CpG sites with a suggestive p-value of <1×10^-5^ were included in the DMR analysis. Results were corrected for multiple testing using the false discovery rate (FDR).

#### Gene Ontology Overrepresentation Analysis

Differentially methylated positions (DMPs) and differentially methylated regions (DMRs) were annotated to genes using the manufacturer’s manifest. Then, Gene Ontology Overrepresentation Analysis (GO ORA) was conducted at the gene level using clusterProfiler. (version 4.14.6)^62^. Additionally, a semantic clustering analysis was performed to identify clusters of related GO terms.

#### GWAS Enrichment Analysis

For GWAS enrichment analysis, we formed three gene sets. The first included all genes associated with DMPs (81 genes), the second reduced these CpG sites to those known to be influenced by genotype^26^ (45 genes), and the third included those associated with the DMRs (81 genes). We then utilized MAGMA (Multi-marker Analysis of GenoMic Annotation)^63^ to test the enrichment of these gene sets in the summary statistics of a recent trans-ancestry meta-analysis of problematic alcohol use and AUD in more than one million participants^27^. To investigate whether genetic variation in smoking behavior-associated genes was driving the results, we also performed a GWAS enrichment analysis for a GWAS of cigarettes per day^30^. In addition, we investigated the association with comorbid disorders, such as major depressive disorder^31^ and posttraumatic stress disorder^32^.

#### Overlap with Alcohol Consumption EWAS

To determine whether the epigenome-wide significant CpG sites identified in the primary analysis were influenced by alcohol consumption, we examined the overlap with the findings of a recent EWAS of alcohol consumption involving more than 8,000 individuals from the Generation Scotland cohort^33^. Fisher’s exact test was used to determine the statistical significance of the overlap.

#### Blood-brain concordance

We further investigated the blood-brain concordance of CpG sites identified in the primary meta-analysis. In the first step, we used BECon^34^ to determine the blood-brain correlations of the top 100 identified CpG sites. In addition, we performed a “look-up” of suggestive significant CpG sites (*p* < 1×10^-5^)^64^ in two recent EWAS of AUD in the dorsolateral prefrontal cortex/Brodmann Area^35,36^, ventral striatum/nucleus accumbens^35,36^, caudate nucleus^35^, putamen^35^, and anterior cingulate cortex^35^.

#### Sensitivity Analysis – Smoking

In some cohorts, a substantial overlap between smoking and AUD was observed (e.g., CIMH). Therefore, we performed sensitivity analyses separately for currently self-reported nonsmoking participants in all cohorts with at least 30 individuals for AUD cases and 30 for controls. The non-smoking meta-analysis consisted of 939 individuals from four cohorts: NIAAA (N=369), UCSF (N=108), NESDA (N=377), and GSMS (N=85). Effect sizes from this meta-analysis were associated with those of the primary meta-analysis using Pearson correlation.

### Validation Cohort

DNA methylation data from the LURIC (Ludwigshafen Risk and Cardiovascular Health Study) was used to replicate methylation risk scores (MRS) from the meta-analysis in an independent cohort. The LURIC study comprises 3,316 individuals hospitalized for coronary angiography between 1997 and 2000^37^. DNA methylation was assayed using the Illumina EPIC BeadChip v1, and the same quality control procedures for array-based data in the discovery analysis were applied, details can be found in Supplementary Text S2. After quality control, data was available for 2,534 individuals^65^. All data presented in this manuscript refers to the subsample with DNA methylation data.

The mean age in the LURIC cohort was M=62.85 years (SD=10.6), and 30.7% of participants were female (N=780). Alcohol consumption was assessed by self-report and converted into grams of ethanol per day^66^. The mean alcohol consumption in grams per day was M=16.61, SD = 24.46, while the median ethanol intake per day was Md=3.08. Overall, the LURIC cohort is characterized by a high percentage of heavy drinkers, with 939 (37.05%) meeting heavy drinking criteria as proposed by the National Institute on Alcohol Abuse and Alcoholism (NIAAA)^39^. For males, the heavy drinking threshold is drinking more than five drinks per day (70 g) or more than 210 g per week. Females fulfill heavy drinking criteria if they consume more than four drinks per day (56 g) or more than 112 g per week. Heavy drinking is considered a risk factor for AUD, which was not assessed in the LURIC cohort.

### Methylation Risk Score

We calculated methylation risk scores (MRS) using the pruning and thresholding approach suggested by Chen et al. (2023)^40^. Here, regional associations between the methylation values of the validation cohort were identified using the CoMeBack algorithm^65^. During the construction of the MRS, one CpG site per region is included in the score, similar to clumping in polygenic risk scores. The minimum p-value and resulting p-value thresholds were constructed from the summary statistics of the meta-analysis, resulting in a minimum p-value of 5.12×10^-17^, representing the p-value of the strongest association in the primary meta-analysis. We tested sixteen thresholds in total ranging from 0.05 to 5×10^-17^ in increments of one decimal point.

To test whether the MRS of AUD was predictive of alcohol consumption and heavy drinking in the LURIC cohort, we constructed a linear model predicting the ethanol intake per day by the MRS while controlling for sex, age, and cell type composition, as described for the primary meta-analysis. All numerical covariates were scaled to the mean. In addition, we tested whether the AUD MRS was predictive of heavy drinking, according to NIAAA criteria, in a generalized linear model using the same covariates as described above. Here, we calculated Nagelkerkes R^2^ to determine the goodness of fit, as well as the area under the receiver operating characteristic curve, using the R package pROC (v. 1.18.5)^67^.

## Supporting information

ExtendedData1

ExtendedData2

ExtendedData3

ExtendedData4

ExtendedData5

## Acknowledgment

We thank all participants in the studies supporting this report.

This work was funded by the Deutsche Forschungsgemeinschaft (DFG, German Research Foundation) – 551108212 to L.Z. and supported by the Intramural Research Program of the National Institutes of Health (NIH) as part of the Division of Intramural Clinical and Biological Research of the National Institute on Alcohol Abuse and Alcoholism (NIAAA) (ZIA-AA000242 to F.W.L). The contributions of the NIH author(s) are considered Works of the United States Government. The findings and conclusions presented in this paper are those of the author(s) and do not necessarily reflect the views of the NIH or the U.S. Department of Health and Human Services.

## Conflicts of Interest

H.R.K. is a member of advisory boards for Altimmune and Clearmind Medicine; a consultant to Sobrera Pharmaceuticals, Altimmune, and Lilly; the recipient of research funding and medication supplies for an investigator-initiated study from Alkermes and a company-initiated study by Altimmune; and an inventor on U.S. provisional patent “Multi-ancestry Genome-wide Association Meta-analysis of Buprenorphine Treatment Response.”

J.M.O. is a current employee and stockholder of Regeneron Pharmaceuticals. Dr. Otto contributed to the manuscript as an analyst while affiliated with the University of Missouri.

M.E.K. and W.M. are employees of Synlab Holding Deutschland GmbH.

K.D. has been a member of the Janssen-Cilag GmbH Steering Committee Neurosciences and received speaker’s honoraria from Janssen-Cilag GmbH until 2022. Currently, K.D. is a member of the Neurotorium Editorial Board, Lundbeck Foundation.

All other authors report no conflicts of interest.

## Data Availability

Full summary statistics are available via figshare:

https://doi.org/10.6084/m9.figshare.30000022.

