## Supplementary figures and images for "DNA Methylation Signatures of Alcohol Use Disorder – A large-scale Meta-Analysis in the Psychiatric Genomics Consortium"

### ExtendedData1

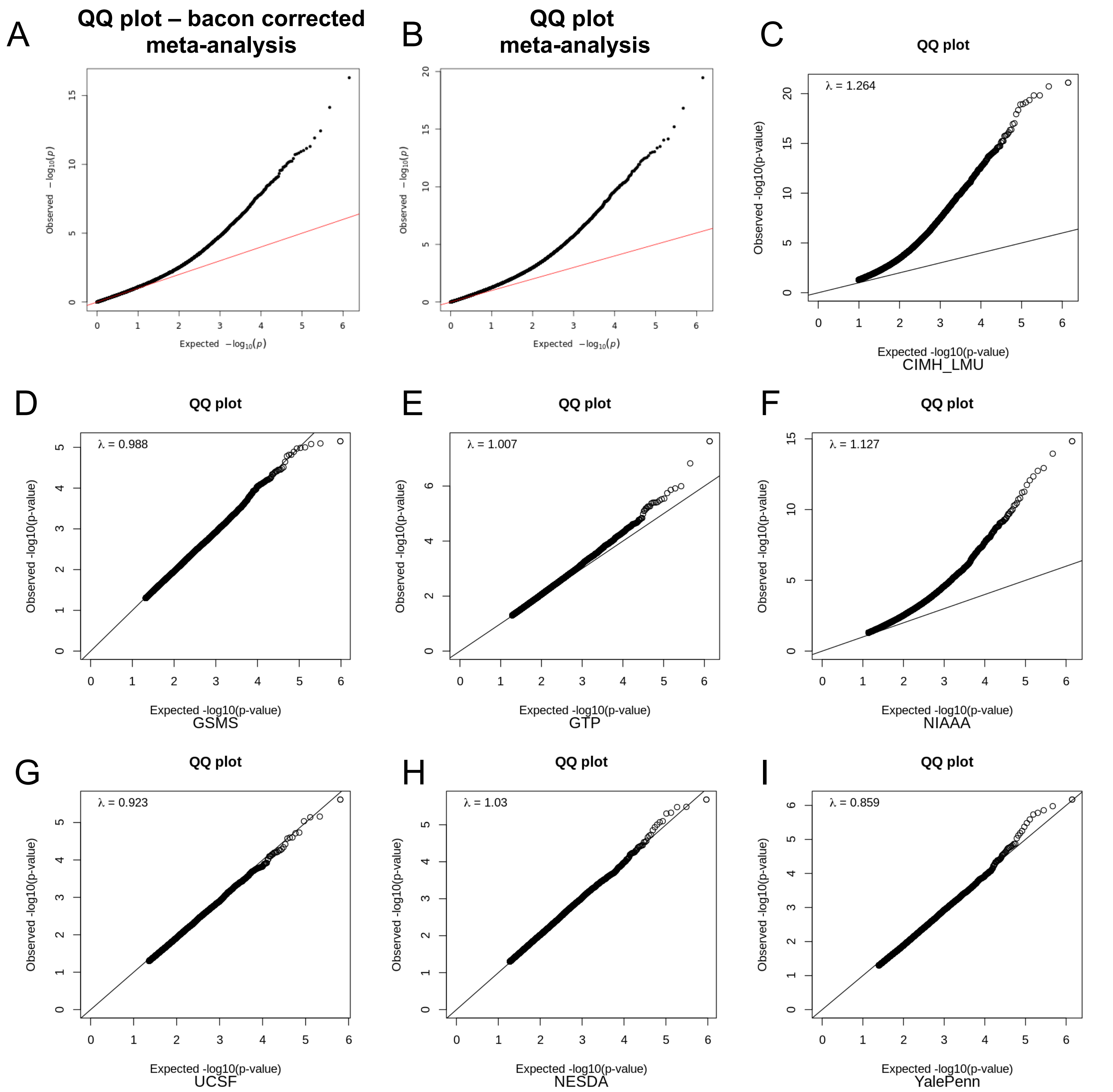

### ExtendedData2

#### Sensitivity Analysis: EPIC & MBD-seq

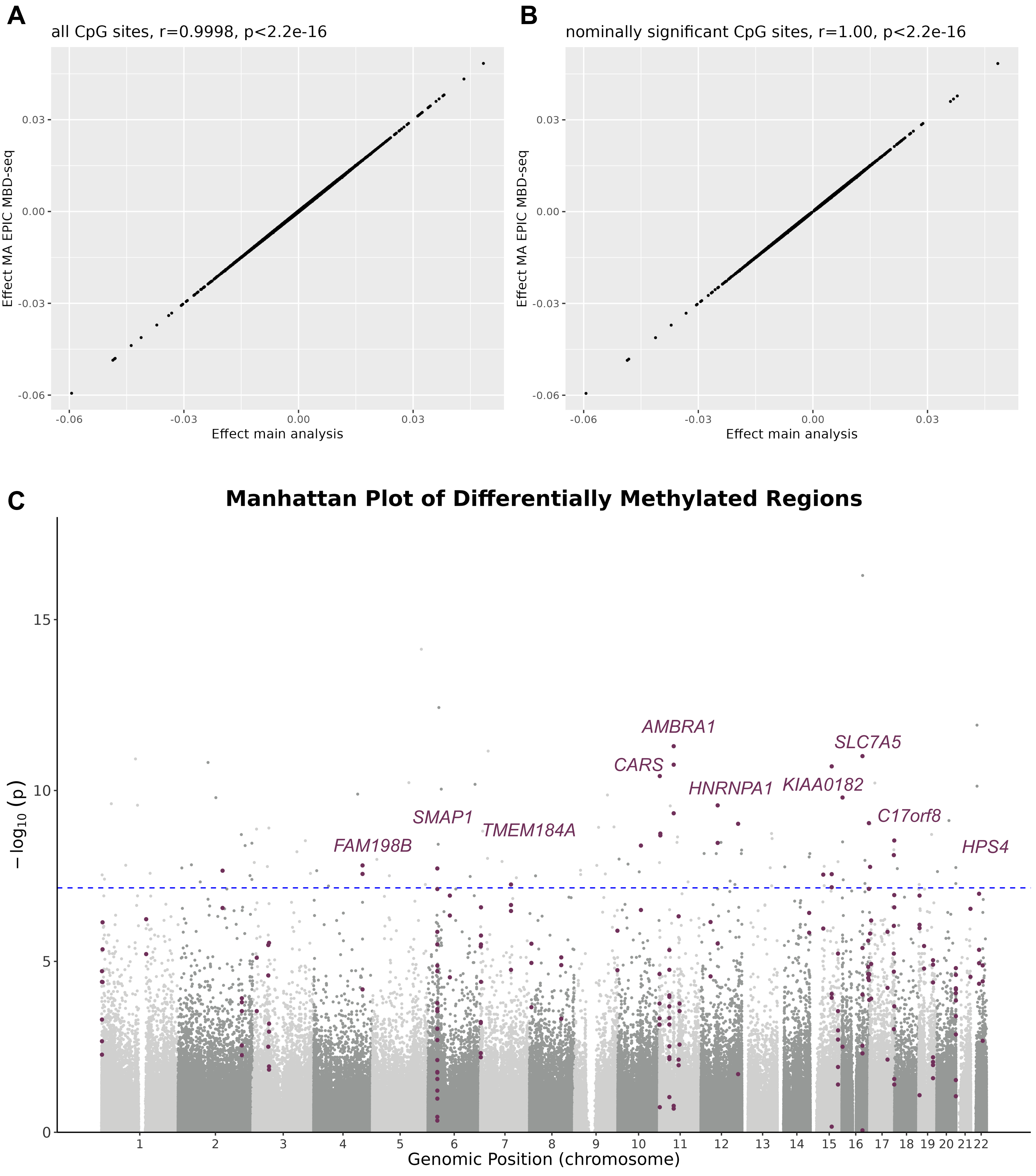

### ExtendedData3

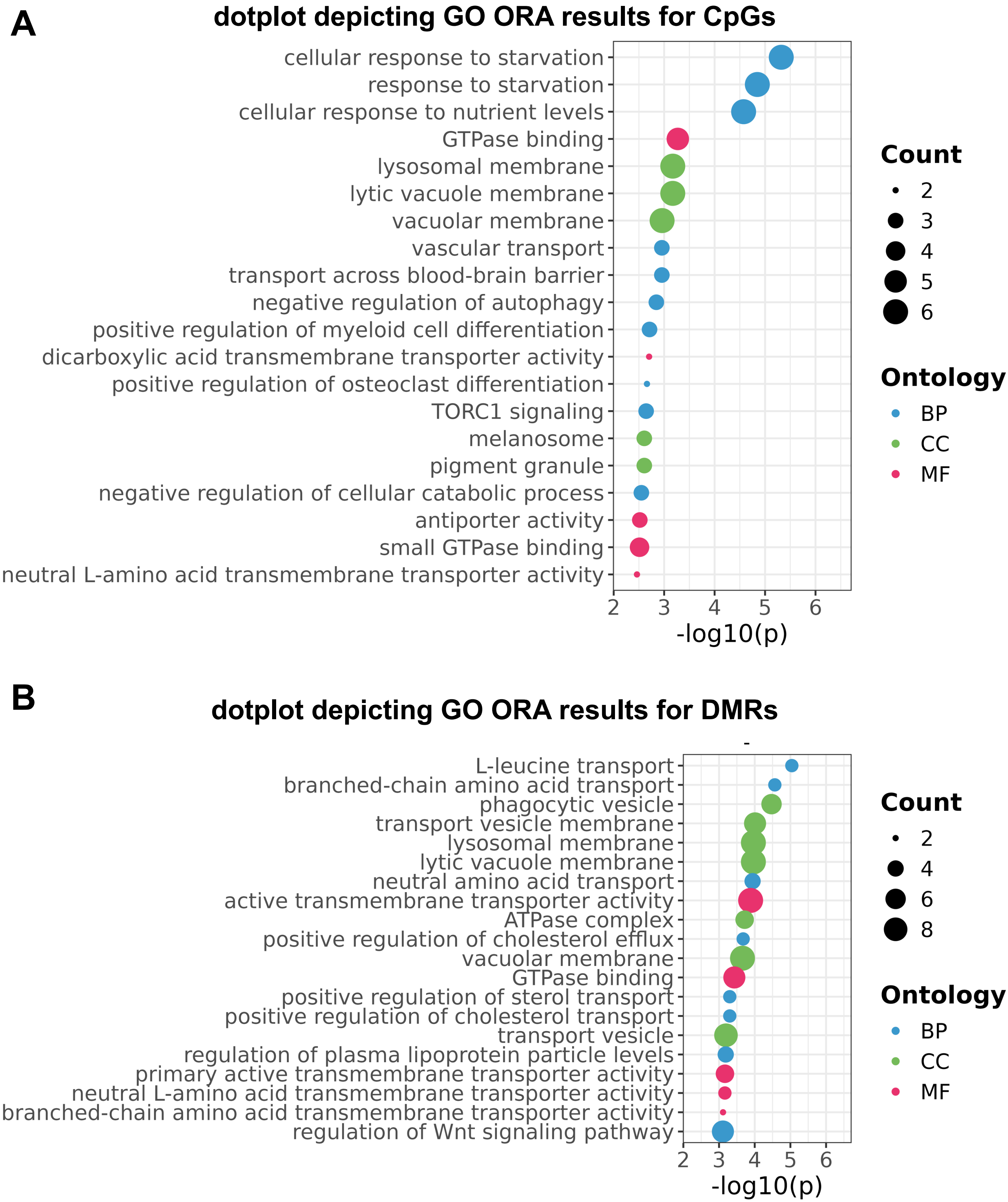

### ExtendedData5

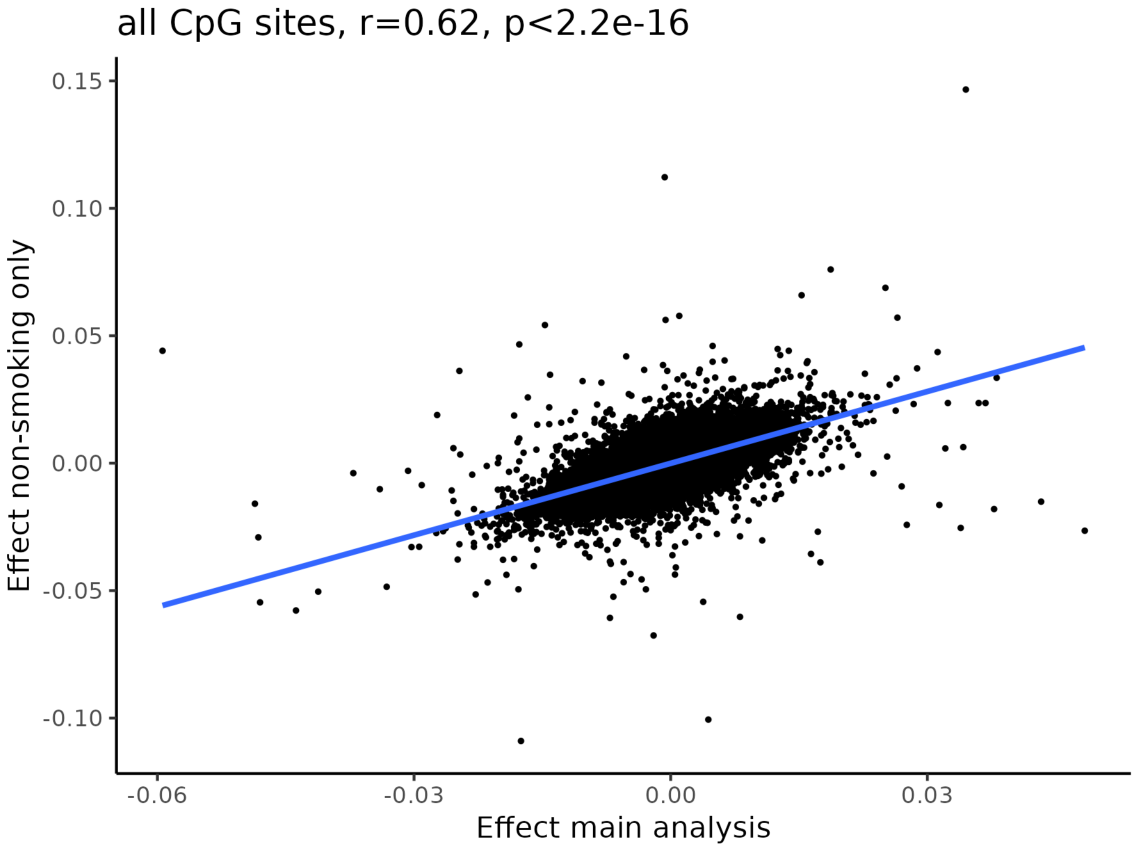

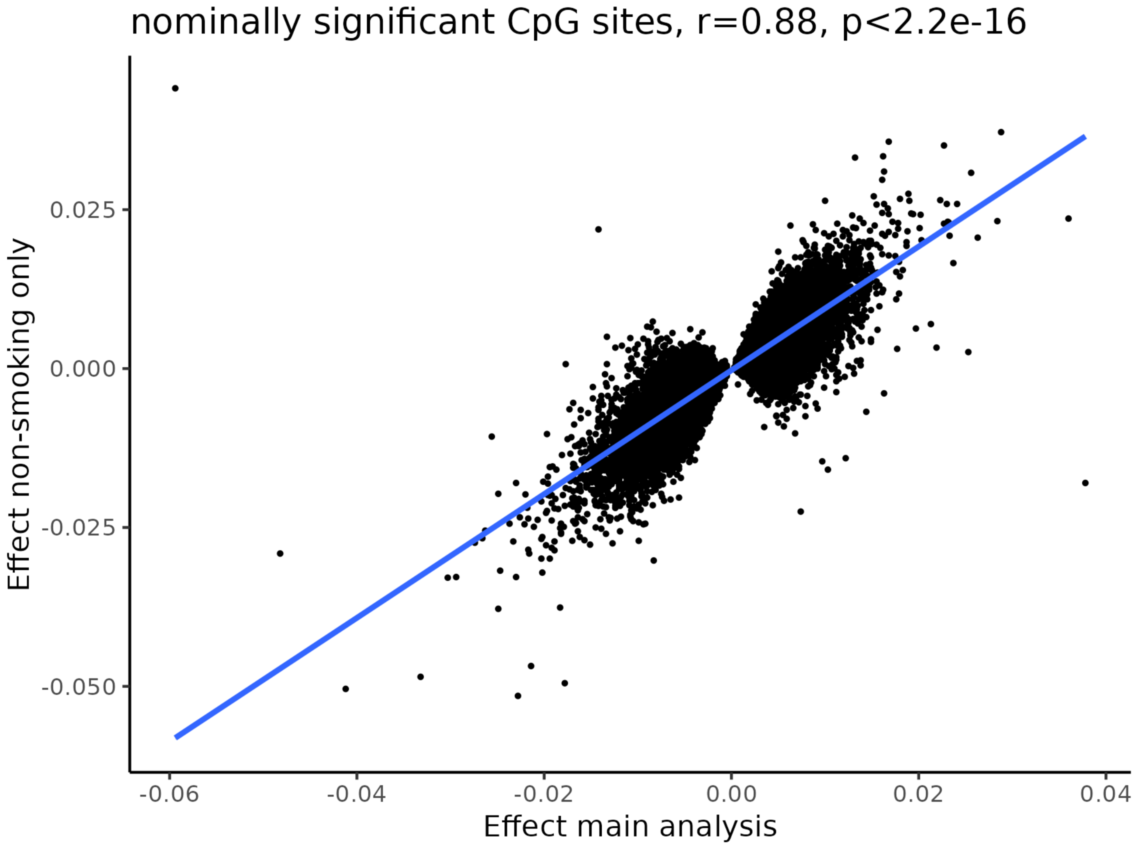
