## ExtendedData4 for "DNA Methylation Signatures of Alcohol Use Disorder – A large-scale Meta-Analysis in the Psychiatric Genomics Consortium"

| CpG | Chr | Coor | Gene(s) | Gene Region(s) | Variability BA10 | Variability BA20 | Variability BA7 | Variability Blood | Correlation BA10 | Correlation BA20 | Correlation BA7 | Cell Composition Blood | Cell Composition Brain |
|---|---|---|---|---|---|---|---|---|---|---|---|---|---|
| cg05603985 | 1 | 2161049 | SKI | intragenic | 0.11 | 0.05 | 0.03 | 0.04 | 0.12 | 0.22 | −0.14 | 0 | 0.02 |
| cg09127607 | 10 | 69914213 | MYPN | intragenic | 0.19 | 0.12 | 0.14 | 0.11 | −0.21 | −0.36 | −0.07 | 0.02 | 0.05 |
| cg11095027 | 11 | 1297066 | TOLLIP | intragenic | 0.17 | 0.11 | 0.11 | 0.09 | −0.24 | 0.36 | 0.33 | 0.02 | 0.04 |
| cg23449274 | 11 | 46564463 | AMBRA1 | intragenic | 0.03 | 0.03 | 0.02 | 0.04 | −0.1 | 0.02 | −0.08 | 0.01 | 0 |
| cg16371518 | 11 | 46564493 | AMBRA1 | intragenic | 0.02 | 0.02 | 0.02 | 0.05 | −0.34 | 0.16 | 0.3 | 0.02 | 0 |
| cg11376147 | 11 | 57261198 | SLC43A1 | intragenic | 0.04 | 0.04 | 0.04 | 0.08 | −0.15 | −0.05 | −0.04 | 0.02 | 0 |
| cg02583484 | 12 | 54677008 | HNRNPA1, HNRNPA1P10 | intragenic, intragenic | 0.06 | 0.05 | 0.06 | 0.1 | 0 | −0.05 | −0.04 | 0.02 | 0 |
| cg14380013 | 12 | 121146751 | UNC119B | promoter | 0.09 | 0.05 | 0.06 | 0.09 | −0.03 | 0.51 | −0.12 | 0.03 | 0.01 |
| cg26829071 | 12 | 131590596 | GPR133 | intragenic | 0.08 | 0.05 | 0.06 | 0.14 | −0.29 | 0.03 | −0.15 | 0.04 | 0.01 |
| cg09940677 | 14 | 103415458 | CDC42BPB | intragenic | 0.24 | 0.16 | 0.15 | 0.1 | −0.32 | −0.19 | 0.34 | 0.01 | 0.06 |
| cg07452706 | 14 | 105649097 | NUDT14 | promoter | 0.1 | 0.06 | 0.05 | 0.15 | −0.21 | −0.22 | 0.11 | 0.04 | 0.02 |
| cg18059012 | 15 | 44969129 | PATL2 | promoter | 0.09 | 0.09 | 0.08 | 0.22 | 0.4 | 0.43 | 0.6 | 0.07 | 0.02 |
| cg17301216 | 15 | 89920348 | LINC00925 | promoter | 0.33 | 0.19 | 0.17 | 0.15 | 0.18 | −0.28 | 0.15 | 0.01 | 0.08 |
| cg06846976 | 16 | 2121682 | TSC2 | intragenic | 0.03 | 0.02 | 0.04 | 0.1 | 0.56 | −0.24 | 0.09 | 0.03 | 0 |
| cg08761535 | 16 | 75079000 | ZNRF1 | intragenic | 0.28 | 0.09 | 0.17 | 0.13 | −0.1 | 0.03 | −0.19 | 0.03 | 0.06 |
| cg07021906 | 16 | 87866833 | SLC7A5 | intragenic | 0.12 | 0.12 | 0.09 | 0.16 | 0.3 | 0.23 | −0.01 | 0.05 | 0.02 |
| cg14330293 | 17 | 1374051 | MYO1C | intragenic | 0.05 | 0.08 | 0.06 | 0.14 | −0.15 | −0.43 | −0.34 | 0.04 | 0.01 |
| cg19379721 | 19 | 19737143 | LPAR2 | intragenic | 0.04 | 0.05 | 0.02 | 0.09 | −0.13 | −0.36 | −0.03 | 0.02 | 0 |
| cg04406114 | 19 | 42599489 | POU2F2 | intragenic | 0.06 | 0.05 | 0.05 | 0.12 | 0.52 | 0.49 | −0.01 | 0.04 | 0.01 |
| cg14817906 | 2 | 97466833 | CNNM4 | intragenic | 0.05 | 0.09 | 0.06 | 0.16 | 0.25 | 0.09 | 0.29 | 0.05 | 0.01 |
| cg11550064 | 2 | 240148191 | HDAC4 | intragenic | 0.24 | 0.17 | 0.11 | 0.13 | −0.41 | −0.21 | −0.37 | 0.04 | 0.05 |
| cg19841423 | 20 | 62366755 | ZGPAT, LIME1 | intragenic, promoter | 0.12 | 0.14 | 0.1 | 0.15 | −0.16 | −0.36 | −0.12 | 0.04 | 0.03 |
| cg03450635 | 4 | 159094213 | FAM198B | promoter | 0.2 | 0.1 | 0.06 | 0.13 | 0.39 | −0.05 | 0.13 | 0.01 | 0.04 |
| cg03996539 | 4 | 159094517 | FAM198B | promoter | 0.23 | 0.13 | 0.11 | 0.11 | 0.3 | −0.16 | 0.22 | 0.02 | 0.04 |
| cg18394552 | 5 | 159428643 | None | intergenic | 0.03 | 0.06 | 0.03 | 0.15 | 0.16 | −0.03 | −0.05 | 0.04 | 0 |
| cg22907189 | 5 | 172430563 | ATP6V0E1 | intragenic | 0.09 | 0.1 | 0.06 | 0.1 | −0.22 | −0.11 | 0.06 | 0.03 | 0.03 |
| cg01538969 | 6 | 30624636 | DHX16 | intragenic | 0.08 | 0.06 | 0.07 | 0.26 | −0.12 | −0.45 | −0.21 | 0.08 | 0 |
| cg17840178 | 6 | 30709803 | FLOT1 | intragenic | 0.2 | 0.08 | 0.09 | 0.13 | −0.01 | −0.01 | −0.08 | 0.04 | 0.04 |
| cg03546163 | 6 | 35654363 | FKBP5 | intragenic | 0.04 | 0.03 | 0.05 | 0.23 | −0.26 | 0.49 | −0.05 | 0.04 | 0 |
| cg11523799 | 6 | 43479817 | YIPF3, None | three_plus, intergenic | 0.17 | 0.1 | 0.09 | 0.13 | −0.08 | 0.34 | 0.04 | 0.05 | 0.03 |
| cg22407942 | 7 | 22894828 | SNORD93 | promoter | 0.07 | 0.06 | 0.04 | 0.13 | −0.05 | −0.17 | −0.41 | 0.02 | 0.01 |
| cg00834988 | 7 | 70060153 | AUTS2 | intragenic | 0.04 | 0.04 | 0.05 | 0.3 | 0.02 | −0.33 | −0.29 | 0.1 | 0.01 |
| cg21227325 | 9 | 78776599 | PCSK5 | intragenic | 0.08 | 0.07 | 0.07 | 0.23 | −0.49 | −0.29 | −0.04 | 0.08 | 0 |

Correlation or Cell Composition Percentile or Variability Status

- 90% (Positive)
- 75−90% (Positive)
- 50−75% (Positive)
- <50% (Positive)
- <50% (Negative)
- 50−75% (Negative)
- 75−90% (Negative)
- 90% (Negative)
- 90% (Blood Cell Comp.)
- 75−90% (Blood Cell Comp.)
- 50−75% (Blood Cell Comp.)
- <50% (Blood Cell Comp.)
- 90% (Brain Cell Comp.)
- 75−90% (Brain Cell Comp.)
- 50−75% (Brain Cell Comp.)
- <50% (Brain Cell Comp.)
- Not Variable
- Variable
- Genomic Info
