## Supplementary Texts for "DNA Methylation Signatures of Alcohol Use Disorder – A large-scale Meta-Analysis in the Psychiatric Genomics Consortium"

### Supplementary Text S1. Cohort Descriptions

#### a. CIMH

**Cohort profile**: Witt et al. (2020)^1^ <https://pubmed.ncbi.nlm.nih.gov/32080920/>

**Design and study population**: The Central Institute of Mental Health (CIMH) Alcohol withdrawal cohort was recruited during a longitudinal study at the CIMH in Mannheim, Germany. For patients, inclusion criteria required a diagnosis of alcohol dependence syndrome according to DSM-IV criteria, as well as the presence of alcohol withdrawal symptoms based on the Clinical Institute Withdrawal Assessment for Alcohol–Revised (CIWA-Ar). Only individuals with a CIWA-Ar score greater than 4 were eligible for inclusion. Exclusion criteria for patients included the presence of any severe mental illness (other than alcohol or nicotine dependence) requiring pharmacological treatment, any severe physical condition that would preclude study participation, inability to provide written informed consent, dependence on substances other than alcohol or nicotine (as defined by DSM-IV), and known neurological disorders. For healthy control participants, in addition to the above exclusion criteria, individuals were excluded if they had any current or past mental disorder (other than nicotine dependence), as assessed by the Structured Clinical Interview for DSM Disorders (SCID), engaged in risky alcohol use—defined as regular consumption exceeding 60 g/day for men or 48 g/day for women, based on the Form90 interview—or had a positive drug screening result.

**Consent and ethical approval**: Written informed consent was obtained from all participants. Ethical approval for the study was obtained at the ethics committee II of the Medical Faculty Mannheim, Heidelberg University. All study procedures were carried out in accordance with the Declaration of Helsinki.

**Division into subcohorts**: None

**Study funding**: This work was supported by the German Federal Ministry of Education and Research (BMBF) through grants Sysmed and SysmedSUD (01ZX1311A, 01ZX1314G, 01EW1810 and 01ZX1311A) within the e:Med research program^2^, and ‘AERIAL’ EE1406C within the Research Net of Psychiatric Diseases, by ERA-NET NEURON grants ‘EMBED’ (01ZX1909A) and ‘PsiAlc’ (01EW1908), by the German Research Foundation grants RI908/11–2 and WI3439/3–2 and the collaborative research center TRR265^3^.

**Acknowledgements**: None

#### b. Yale-Penn

**Cohort profile**:
Genetics: Gelernter et al. (2014)^4^ <https://pubmed.ncbi.nlm.nih.gov/24143882/>
Epigenetic: Pathak et al. (2025)^5^ <https://pubmed.ncbi.nlm.nih.gov/40247127/>

**Design and study population**: Yale-Penn participants were recruited from five U.S. medical centers, including the Yale University School of Medicine (APT Foundation, New Haven, CT) and the University of Pennsylvania Perelman School of Medicine (Philadelphia, PA). All participants were evaluated using the Semi-Structured Assessment for Drug Dependence and Alcoholism (SSADDA), which captures diagnoses of Alcohol Use Disorder (AUD), other Substance Use Disorders (SUDs), and psychiatric conditions according to DSM-IV criteria. DNA methylation data were available for 886 individuals, generated using the Infinium® MethylationEPIC BeadChip array (Illumina, San Diego, CA). Two models were tested in this cohort: the first restricted the analysis to individuals with AUD and no other SUDs (n = 247), while the second included individuals with both AUD and SUDs, adjusting for SUD diagnoses as covariates alongside age and sex.

**Consent and ethical approval**: Informed consent was signed for all participants and approved by the institutional review board at each site. Confidentiality certificates were obtained from the National Institute on Drug Abuse and the National Institute on Alcohol Abuse and Alcoholism.

**Division into subcohorts**: Yes, based on the PI who sponsored sample analysis.

**Study funding**: R.P. acknowledges grants from the National Institute of Mental Health (RF1 MH132337) and One Mind Rising Star Award. D.F.L. is funded by a Career Development Award from the US Department of Veterans Affairs Office of Research and Development (1IK2BX005058). J.L.M.O. acknowledges support from U.S. Department of Veterans Affairs via 1IK2CX002095 and NIDA R21 DA050160. J.G. reports support from the Department of Veterans Affairs (5IO1CX001849-04 and the VISN 1 New England MIRECC) and NIH/NIDA (R01 DA037974, R01 DA058862).

**Acknowledgements:** None

#### c. UCSF

**Cohort profile**: Vieten et al. (2004)^6^ <https://pubmed.ncbi.nlm.nih.gov/15597083/>

**Design and study population**: The University of California San Francisco Family Alcoholism Study is a nationwide study on the genetics of alcoholism and other substance dependence. Probands were sampled from the community through semi-targeted direct mail, a web site, press releases, advertisements and from alumni of treatment centers across the nation. Probands were invited to participate if they met screening criteria for alcohol dependence at some point in their lifetime and had at least one sibling or both parents available to participate in the study. With the permission of the proband, relatives were invited by mail to participate. A modified version of the Semi-Structured Assessment for the Genetics of Alcoholism was used to make DSM-IV alcohol and other substance misuse and psychiatric diagnoses. Probands with stimulant, cocaine, or opioid dependence and those who reported any history of intravenous substance use were excluded. Probands were also excluded if, upon screening, they reported a current or past diagnosis of schizophrenia, bipolar disorder, or other psychiatric illness involving psychotic symptoms, a life-threatening illness, or an inability to speak and read English.

**Consent and ethical approval**: Institutional review board committees approved all data collection, and participants provided informed consent prior to participation.

**Division into subcohorts**: None.

**Study funding**: This work was supported by grants from the National Institute of Alcohol Abuse and Alcoholism: F31AA025269 (principal investigator: Jacqueline M. Otto) and the National Institute on Drug Abuse: R01DA030976 (principal investigators: Kirk C. Wilhelmsen, Ian R. Gizer), and the State of California for medical research on alcohol and substance abuse through the University of California at San Francisco and Ernest Gallo Clinic and Research Center to Kirk C. Wilhelmsen.

**Acknowledgements**: None.

#### d. GTP

**Cohort profile**: Gillespie et al. (2009)^7^ <https://pubmed.ncbi.nlm.nih.gov/19892208/>

**Design and study population**: Participants were recruited as part of the Grady Trauma Project (GTP), which investigates the influence of genetic and environmental factors on responses to stressful life events in a predominantly African American, urban population of low socioeconomic status. GTP research participants were approached in the waiting rooms of the primary care clinic or obstetrical-gynecological clinic of a large, urban, public hospital in Atlanta, GA while either waiting for their medical appointments or while waiting with others who were scheduled for medical appointments. Alcohol use disorder (AUD) was assessed by clinical psychologists using the Structured Clinical Interview for DSM-IV Axis I Disorders (SCID-IV) or the Mini International Neuropsychiatric Interview (MINI) version 5 or 7. The alcohol use disorders identification test (AUDIT) was applied in a subset of participants^8^. Demographic variables including age, sex, race, and smoking were assessed through self-report.

**Consent and ethical approval**: Subjects willing to participate provided written informed consent and participated in a verbal interview and blood draw. All procedures in this study were approved by the Institutional Review Board of Emory University School of Medicine and the Grady Health Systems Research Oversight Committee.

**Division into subcohorts**: None.

**Study funding**: GTP is supported by the National Institutes of Mental Health (MH096764 to KJR and MH071537 to CFG&KJR).

**Acknowledgements:** None.

#### e. NIAAA

**Cohort profile**: Lohoff et al. (2021)^9^ <https://pubmed.ncbi.nlm.nih.gov/32398718/>

**Design and study population**:

**Consent and ethical approval**:

**Division into subcohorts**:

**Study funding**:

**Acknowledgements**:

#### f. GSMS

**Cohort profile**: Costello et al. (1996)^10^ <https://pubmed.ncbi.nlm.nih.gov/8956679/>

**Design and study population**: The Great Smoky Mountain Study (GSMS) is an ongoing, prospective longitudinal, representative study of children in 11 predominantly rural counties in the southeast United States, which began in 1993. Three cohorts of children, age 9 to 13 years, were recruited resulting in N=1,420 participants. As part of a larger methylation study on childhood and young adult exposures, 525 participants were selected that were 9-21 years of age and had a bloodspot taken at the time of assessment available. Of the 525 participants, 412 had bloodspots included from older time points (age 18 years or older). These participants were used in the current meta-analyses. Participant substance involvement was assessed during an in-person interview. At each interview assessment the participant completed a full structured clinical interview about substance involvement using the Young Adult Psychiatric Assessment (YAPA). The substance use module of the YAPA included assessments of DSM-IV abuse and dependence. Although DSM-5 alcohol use disorder (AUD) symptoms of craving and withdrawal were not part of the DSM-IV abuse or dependence diagnostic criteria, these data have been collected since the start of GSMS is 1993.

**Consent and ethical approval**: Participants provided informed consent, and the study was approved by the Institutional Review Boards at Duke University and Virginia Commonwealth University.

**Division into subcohorts**: none.

**Study funding**: This study was supported by the National Institutes of Health (R01 R01MH104576 to Edwin J.C.G. van den Oord, R01AA026057 to Shaunna L. Clark & R01AA030116 to Edwin J.C.G. van den Oord and Shaunna L. Clark). The funding sources had no role in the study design, writing of the report or decision to submit the article for publication.

**Acknowledgements**: None.

#### g. NESDA

**Cohort profile**: Penninx et al. (2008)^11^ <https://pubmed.ncbi.nlm.nih.gov/18763692/>

**Design and study population**: The Netherlands Study of Depression and Anxiety (NESDA) is an ongoing longitudinal multi-center, cohort study designed to investigate the long-term course and consequences of depression and anxiety disorders. Its 2,981 participants include patients with a current or lifetime diagnosis of depression and/or anxiety disorder and controls without any lifetime depressive disorder and /or anxiety disorder. As part of a larger project on methylation and depression, a sample of 1,132 participants with blood samples available at study entry were selected for methylation sequencing. Lifetime AUD was diagnosed using the DSM-IV based Composite International Diagnostic Interview (CIDI version 2.1)^12^ that was administered by specially trained research staff. AUD cases included individuals that met criteria for either abuse or dependence on the DSM-IV. Controls had initiated alcohol use but did not qualify for an AUD diagnosis.

**Consent and ethical approval**: All participants provided written informed consent. The study was approved by ethical committees at all participating centers in the Netherlands and the Institutional Review Board at Virginia Commonwealth University.

**Division into subcohorts**: None.

**Study funding**: The infrastructure for the NESDA study (www.nesda.nl) is funded through the Geestkracht program of the Netherlands Organisation for Health Research and Development (ZonMw, grant number 10-000-1002) and financial contributions by participating universities and mental health care organizations (VU University Medical Center, GGZ inGeest, Leiden University Medical Center, Leiden University, GGZ Rivierduinen, University Medical Center Groningen, University of Groningen, Lentis, GGZ Friesland, GGZ Drenthe, Rob Giel Onderzoekscentrum). This methylation data generation and AUD analyses were supported by the National Institutes of Health (R01MH099110 to Edwin J.C.G. van den Oord, R01AA026057 to Shaunna L. Clark & R01AA030116 to Edwin J.C.G. van den Oord and Shaunna L. Clark). The funding sources had no role in the study design, writing of the report or decision to submit the article for publication.

**Acknowledgements**: None.

#### h. LURIC

**Cohort profile:** Winkelmann et al. (2001). ^13^<https://pubmed.ncbi.nlm.nih.gov/11258203/>

**Design and study population**: LURIC study is a monocentric hospital-based study that recruited 3316 participants of European ancestry that had been referred for coronary angiography between 1997 and 2000 at the Ludwigshafen Heart Center in South-West Germany. Clinical indications for angiography were chest pain or a positive non-invasive stress test suggestive of myocardial ischemia. Individuals suffering from acute illnesses other than acute coronary syndrome, chronic non-cardiac diseases and a history of malignancy within the past five years were excluded.

**Consent and ethical approval**: The ethics committee of the "Landesärztekammer Rheinland-Pfalz" approved the study (837.255.97 (1394)) that was conducted in accordance with the "Declaration of Helsinki”. Informed written consent was obtained from all participants.

**Division into subcohorts:** None

**Study funding:** The DNA methylation measurement in LURIC was supported by the 7th Framework Program RiskyCAD (grant agreement number 305739) of the European Union and by the German Ministry for Education and Research as part of the Competence Cluster for Nutrition and Cardiovascular Health (nutriCARD) Halle-Jena-Leipzig (grant agreement numbers 01EA1411A and 01EA1808A)

**Acknowledgements:** We thank all participants of the LURIC study as well as the study team who were either temporarily or permanently involved in patient recruitment as well as sample and data handling, in addition to the laboratory staff at the Ludwigshafen General Hospital and the University Hospitals of Freiburg, Ulm, Mannheim, Germany, and Graz, Austria.

### Supplementary Text S2. Quality Control of DNA methylation data in the LURIC cohort.

Quality Control in the LURIC cohort was conducted similarly to the main analysis. Raw IDAT files were processed using the *minfi* package, and Illumina background subtraction was applied to eliminate outliers by averaging the negative control signal intensity. To address technical biases in the EWAS, a principal component analysis (PCA) was performed on the positive control probe intensities. Probes were excluded if their detection p-value exceeded 0.001, resulting in a call rate below 95%, if the bead count was lower than three in over 5% of samples, or if they were absent from the autosomes. Samples were discarded if their call rate fell below 95% or if there was a discrepancy between reported and biological sex. The EpiDISH package was utilized to estimate cell type composition, yielding estimates for CD8 T cells, CD4 T cells, natural killer cells (NK), B cells (B), monocytes (mono), and neutrophils.

1	Witt, S. H. *et al.* Acute alcohol withdrawal and recovery in men lead to profound changes in DNA methylation profiles: a longitudinal clinical study. *Addiction* **115**, 2034-2044, doi:10.1111/add.15020 (2020).

2	Spanagel, R. *et al.* A systems medicine research approach for studying alcohol addiction. *Addict Biol* **18**, 883-896, doi:10.1111/adb.12109 (2013).

3	Heinz, A. *et al.* Addiction Research Consortium: Losing and regaining control over drug intake (ReCoDe)-From trajectories to mechanisms and interventions. *Addict Biol* **25**, e12866, doi:10.1111/adb.12866 (2020).

4	Gelernter, J. *et al.* Genome-wide association study of opioid dependence: multiple associations mapped to calcium and potassium pathways. *Biol Psychiatry* **76**, 66-74, doi:10.1016/j.biopsych.2013.08.034 (2014).

5	Pathak, G. A. *et al.* Epigenetic and genetic profiling of comorbidity patterns among substance dependence diagnoses. *Mol Psychiatry*, doi:10.1038/s41380-025-03031-y (2025).

6	Vieten, C., Seaton, K. L., Feiler, H. S. & Wilhelmsen, K. C. The University of California, San Francisco Family Alcoholism Study. I. Design, methods, and demographics. *Alcohol Clin Exp Res* **28**, 1509-1516, doi:10.1097/01.alc.0000142261.32980.64 (2004).

7	Gillespie, C. F. *et al.* Trauma exposure and stress-related disorders in inner city primary care patients. *Gen Hosp Psychiatry* **31**, 505-514, doi:10.1016/j.genhosppsych.2009.05.003 (2009).

8	Almli, L. M. *et al.* Problematic alcohol use associates with sodium channel and clathrin linker 1 (SCLT1) in trauma-exposed populations. *Addict Biol* **23**, 1145-1159, doi:10.1111/adb.12569 (2018).

9	Lohoff, F. W. *et al.* Epigenome-wide association study and multi-tissue replication of individuals with alcohol use disorder: evidence for abnormal glucocorticoid signaling pathway gene regulation. *Mol Psychiatry* **26**, 2224-2237, doi:10.1038/s41380-020-0734-4 (2021).

10	Costello, E. J. *et al.* The Great Smoky Mountains Study of Youth. Goals, design, methods, and the prevalence of DSM-III-R disorders. *Arch Gen Psychiatry* **53**, 1129-1136, doi:10.1001/archpsyc.1996.01830120067012 (1996).

11	Penninx, B. W. *et al.* The Netherlands Study of Depression and Anxiety (NESDA): rationale, objectives and methods. *Int J Methods Psychiatr Res* **17**, 121-140, doi:10.1002/mpr.256 (2008).

12	Wittchen, H. U. Reliability and validity studies of the WHO--Composite International Diagnostic Interview (CIDI): a critical review. *J Psychiatr Res* **28**, 57-84, doi:10.1016/0022-3956(94)90036-1 (1994).

13	Winkelmann, B. R. *et al.* Rationale and design of the LURIC study--a resource for functional genomics, pharmacogenomics and long-term prognosis of cardiovascular disease. *Pharmacogenomics* **2**, S1-73, doi:10.1517/14622416.2.1.S1 (2001).
